# An integrated polygenic and clinical risk tool enhances coronary artery disease prediction

**DOI:** 10.1101/2020.06.01.20119297

**Authors:** Fernando Riveros-Mckay, Michael E. Weale, Rachel Moore, Saskia Selzam, Eva Krapohl, R. Michael Sivley, William A. Tarran, Peter Sørensen, Alexander S. Lachapelle, Jonathan A. Griffiths, Ayden Saffari, John Deanfield, Chris C. A. Spencer, Julia Hippisley-Cox, David J. Hunter, Jack W O’Sullivan, Euan A Ashley, Vincent Plagnol, Peter Donnelly

**Author notes:** These authors jointly supervised the work (Vincent Plagnol, Peter Donnelly).

## Abstract

**Background:** There is considerable interest in whether genetic data can be used to improve standard cardiovascular disease risk calculators, as the latter are routinely used in clinical practice to manage preventative treatment.

**Methods:** This research has been conducted using the UK Biobank (UKB) resource. We developed our own polygenic risk score (PRS) for coronary artery disease (CAD), using novel and established methods to combine published genomewide association study (GWAS) data with data from 114,196 UK Biobank individuals, also leveraging a large resource of other GWAS datasets along with functional information, to aid in the identification of causal variants, and thence define weights for > 8M genetic variants. We utilised a further 60,000 UKB individuals to develop an integrated risk tool (IRT) that combined our PRS with established risk tools (either the American Heart Association/American College of Cardiology’s pooled cohort equations (PCE) or the UK’s QRISK3) which was then tested in an additional, independent, set of 212,563 UKB individuals. We evaluated prediction performance in individuals of European ancestry, both as a whole and stratified by age and sex.

**Findings:** The novel CAD PRS showed superior predictive power for CAD events, compared to other published PRSs. As an individual risk factor, it has similar predictive power to each of systolic blood pressure, HDL cholesterol, and LDL cholesterol, but is more predictive than total cholesterol and smoking history. Our novel CAD PRS is largely uncorrelated with PCE, QRISK3, and family history, and, when combined with PCE into an integrated risk tool, had superior predictive accuracy. In individuals reclassified as high risk, CAD event rates were markedly and significantly higher compared to those reclassified as low risk. Overall, 9.7% of incident CAD cases were misclassified as low risk by PCE and correctly classified as high risk by the IRT, in contrast to 3.7% misclassified by the IRT and correctly classified by PCE. The overall net reclassification improvement for the IRT was 5.7% (95% CI 4.4−7.0), but when individuals were stratified into four age-by-sex subgroups the improvement was larger for all subgroups (range 7.7%−17.3%), with best performance in younger middle-aged men aged 40–54yo (17.3%, 95% CI 13.0–21.5). Broadly similar results were found using a different risk tool (QRISK3), and also for cardiovascular disease events defined more broadly.

**Interpretation:** An integrated risk tool that includes polygenic risk outperforms current, clinical risk stratification tools, and offers greater opportunity for early interventions. Given the plummeting costs of genetic tests, future iterations of CAD risk tools would be enhanced with the addition of a person’s polygenic risk.

**Funding:** Genomics plc

## Introduction

Cardiovascular disease (CVD) is a major cause of morbidity and mortality worldwide ^1^. A risk-based prevention strategy, with prevention efforts most strongly targeted towards those at higher risk, is the widely accepted approach to disease prevention ^2^. In the US, the risk prediction algorithm recommended by the American College of Cardiology / American Heart Association is the pooled cohort equations (PCE) algorithm ^3,4^, while in the UK the National Institute for Health and Care Excellence recommends the QRISK algorithm, with the latest version being QRISK3 ^5,6^. Both algorithms predict CVD risk over 10 years, based on multiple risk factors including age, sex, ethnicity, smoking history, systolic blood pressure, cholesterol levels and co-morbidities. Active management, which may include lipid-lowering treatment, is recommended for individuals whose 10-year risk is predicted to be above a certain threshold (7.5% in the US, 10% in the UK).

An important component of cardiovascular disease is coronary artery disease (CAD), which has been a particular focus for genetic studies. Family and GWAS studies have estimated a heritability for CAD of between 40%−60% ^7,8^. More recently, studies have shown that a polygenic risk score (PRS) derived from large-scale genome-wide genotype data can have predictive power for CAD ^9–11^, raising the question of whether it would be beneficial to add PRS to existing risk predictors to aid the identification of high-risk individuals. As it remains constant over the life course, a PRS could be used to guide disease prevention earlier in life before standard risk factors have an appreciable impact.

Three recent studies in middle-aged individuals of European ancestry have combined a PRS for CAD with standard risk prediction algorithms ^12–14^. One large study of 352,660 adults in the UK Biobank found a significant overall net reclassification improvement (NRI) (4.4% against the PCE algorithm, 95% CI 3.1 – 4.9) ^12^, and discrimination when stratifying by age (≥55yo and <55yo). Smaller studies in the ARIC cohort (n = 4,847)^13^, MESA cohort (n = 2,390) ^13^, and FINNRISK cohort (n = 21,813) ^14^ found a non-significant NRI for risk tools that combined PRS and standard risk tools, compared to the standard risk tool alone. The FINNRISK study also examined predictive performance in ≥55yo and <55yo age groups and found a significant NRI in the younger group, but not in the older group.

While some of the above studies examined the predictive accuracy of an integrated PRS and clinical risk tool in different age groups, this stratification was limited (only < or ≥ 55yo) and did not additionally stratify age-groups by sex. Given the uncertainty surrounding the clinical utility of an integrated genetic and clinical cardiovascular risk tool, we set out to definitively address this. With our access to the largest genome-wide association study (GWAS) results, and enhanced methods to construct PRS (using a combination of novel and established methodologies), we examine the clinical utility of an integrated genetic and clinical risk prediction tool both overall, and across a broad array of age-by-sex subgroups.

## Methods

### Study participants

We considered UK Biobank (UKB) individuals of European ancestry (n = 449,474), aged between 40 and 69yo at recruitment (median age = 58.3, interquartile range = 51.2 – 63.6), of which 46% are men. The UKB cohort has been previously described in detail elsewhere ^15,16^. Genotype data were generated using an Affymetrix array as described by Bycroft et al ^16^. We limited the analysis to 449,011 samples that passed genotype imputation quality control and ascribed to European ancestry, as previously described ^16^.

UKB recruitment occurred between 2006–2010. Information on disease status, operations, medications, and lifecourse exposures were obtained from linkage with secondary care hospital records, death records, and through an interview and baseline assessment at recruitment. Biomarker data were obtained from stored serum samples ^17^. Prospective analyses were based on seven years of follow-up, with incident data (post baseline assessment) obtained through secondary care hospital records and death records, available through March 2017.

### Phenotype definitions

To define individuals in UK Biobank with CVD and CAD, we combined information from hospital electronic records, death records, and self-reported conditions and operations at the time of UK Biobank recruitment (as described by Elliott et al ^12^ in their eTable 2). CAD was defined using ICD10 codes I21-I23, I24.1, I25.2; ICD-9 codes 410–412, 429.79; self-report of heart attack or myocardial infarction (UKB code 1075 in field 20002; code 1 in field 6150); or self-report of coronary angioplasty or coronary artery bypass grafts (OPCS-4 codes K40.1–4, K41.14, K45.1–5, K49.1–2, K49.8–9, K50.2, K75.1–4, K75.8–9; UKB codes 1070, 1095 in field 20004). CVD (used for secondary analyses and defining sub-cohorts) was defined as individuals with CAD plus ICD-10 codes G45, I20-I25, I63-I64; ICD-9 codes 413–414, 434, 436; self-report of angina, stroke, or transient ischaemic attack (UKB codes 1074, 1082, 1583 in field 20002; codes 2,3 in field 6150); or self-report of cardiovascular procedures (OPCS-4 codes K40-K46, K47.1, K49-K50, K75; UKB codes 1071, 1105, 1109, 1514 in field 20004).

### Standard risk tools

Calculation of the American College of Cardiology/American Heart Association pooled cohort equations (PCE) 10-year predicted risk for atherosclerotic cardiovascular disease was implemented as described by Goff et al ^3^. The calculation requires knowledge of a person’s age, sex, ethnicity (White / African American / Other), systolic blood pressure, total and high-density lipoprotein cholesterol, and history of diabetes, smoking, and hypertension treatment. Definition of these variables is described in the supplementary appendix (Table S1).

As a secondary analysis, we calculated the QRISK3 10-year predicted CVD risk as described by Hippisley-Cox et al ^5^. The calculation requires knowledge of a wider range of variables than the PCE tool, as described in the supplementary appendix.

### UK Biobank subgroup definitions

We used distinct subsets of UK Biobank in different parts of our analysis. Specifically, we partitioned UKB individuals of European ancestry into four distinct subsets according to their CVD and statin usage status and missing data status for calculating PCE and QRISK3 clinical risk scores (see Figure S1 for a flow chart description):

– Group I, “no PCE/QRISK3 available”, included 114,196 individuals with missing data that prevented PCE or QRISK3 calculation;
– Group II, “training”, included 60,000 randomly sampled European ancestry individuals that were CVD-free and not taking statins at UKB assessment;
– Group III, “test”, included 212,563 individuals of European ancestry that were CVD-free and not taking statins at UKB assessment and were not selected to be part of Group II;
– Group IV, “statin usage and prevalent cardiovascular disease”, included 62,252 individuals with prevalent CVD or taking statins at the time of UKB assessment.

The focus of our study was on the development and comparative evaluation of different prognostic prediction models for CAD, using separate development and test sets (a Type 2 study according TRIPOD guidelines ^18^, with Groups I and II used as development and Groups III and IV as test sets). Group I was used to generate additional GWAS data for CAD, using both prevalent and incident CAD outcomes, to augment pre-existing published CAD GWAS data. Individuals who were free of statin usage, with no prevalent cardiovascular disease, and with sufficient data for the calculation of PCE and QRISK3, were randomly split into Groups II and III. Group II was used to train coefficients and hyperparameters in our models, and Group III was used to test the performance of our models on incident CAD and CVD outcomes (events following UKB baseline assessment). Group IV was used in combination with Group III to test the performance of our models on prevalent CAD and CVD outcomes (events prior to UKB assessment), or on all (prevalent+incident) outcomes. Information on the distribution of follow-up times in each Group is given in the supplementary appendix (Table S2).

### CAD polygenic risk score

We derived a PRS for CAD using a three step approach, combining the largest available published CAD GWAS (Nikpay et al ^19^, 60,801 cases, 123,504 controls) with additional CAD GWAS data from UKB Group I (6,868 cases, 107,282 controls). In the first step, we used a Bayesian non-parametric multi-trait colocalization and fine-mapping procedure (see supplementary appendix) to identify 5,542 variants with good evidence for a causal effect on one or more traits in the Genomics plc repository of > 5,000 GWAS summary statistic datasets, screened to be free of UKB samples. In the second step, we computed the posterior odds ratios for association with CAD at each of these variants, using a fixed effects meta-analysis of the Nikpay et al GWAS with our in-house UKB Group I GWAS, and following LDpred methodology ^20^ restricted to these 5,542 variants only. In the third step, we conditioned the remaining 8,279,162 variants in the meta-analysed CAD GWAS on the posterior odds ratios obtained in the second step, and performed an additional LDpred computation on these variants, adjusting the priors for functionally informative variants following Marquez-Luna et al ^21^. Group II was used to optimise LDpred hyperparameters. The result was a PRS with weights spread over > 8M variants.

We assessed PRS performance using Cox regression, controlling for age at UKB assessment, sex and the first five genetic principal components as computed by Bycroft et al ^16^. The coefficients for this model were trained in Group II, and tested either in Group III (for incident outcomes) or Groups III and IV (for prevalent and prevalent+incident outcomes). CAD outcomes occurring more than 7 years after UKB assessment were right-censored, due to sparsity of information on these events. Following Elliott et al ^12^, we measured PRS effect size using the hazard ratio difference at 1 SD of PRS (HR per SD), and discriminative performance was evaluated using Harrell’s C index, a metric equivalent to the area under the receiver operating characteristic curve, but with accommodation for censored survival data ^22–24^.

### Combination of CAD PRS and PCE into an Integrated Risk Tool

Our integrated risk tool (IRT) calculates the combined odds of a CAD event in the next 10 years. We illustrate with PCE, but the same procedure applies to QRISK3. The IRT multiplies the 10-year odds obtained from the PCE with the odds ratio implied by a person’s CAD PRS. The formula for the odds ratio was obtained from a logistic regression model fitted to incident CAD outcomes in the Group II training set, in which terms for the PCE and PRS main effects and their interaction were fitted. The logistic regression was performed separately on men and women, to accommodate the separate PCE calculations applied to men and women. Further details are given in the supplementary appendix.

Model performance was assessed using observed incident CAD outcomes in Group III, for a 7-year follow-up time after UKB assessment. Reclassification improvement was evaluated using the categorical Net Reclassification Improvement (NRI) metric ^25^, which is the sum of the net reclassification improvement in cases (the proportion of cases correctly reclassified as high risk minus those incorrectly reclassified as low risk) and the net reclassification improvement in noncases (the proportion of noncases correctly reclassified as low risk minus those incorrectly reclassified as high risk). High and low risk categories were defined using 7.5% and 10% 10-year risk thresholds for PCE and QRISK3 respectively. Following Elliott et al ^12^, we present the NRI multiplied by 100 as a pseudo-percentage (range, −200 to 200), and we present both the full NRI metric, as defined above, as well as the within-case and within-noncase subcomponents.

### Age dependent survival analysis

We fitted a generalised survival model for CAD events against a binary indicator variable for individuals in the top 10% of the PRS distribution, allowing for an age-dependent hazard ratio (HR) using the R package Rstpm2 ^26^. This generated the estimated hazard ratio applicable to the top 10% of the individuals according PRS-based CAD risk, compared to the remainder of the cohort. We used Group III for this analysis (incident CAD cases only).

## Results

### CAD PRS performance

We considered first the independent predictive performance of the CAD PRS, both overall and separated into age-by-sex subgroups. Overall, our PRS was a significant predictor of CAD in both prevalent (pre-UKB assessment) and incident (post-UKB assessment) outcomes (prevalent non-events and events obtained from Groups III and IV respectively, Harrell’s C = 0.67, HR per SD increase = 1.82, 95% CI 1.79–1.86, p<10^100^; incident outcomes obtained from Group III, Harrell’s C = 0.64, HR per SD increase = 1.64, 95% CI 1.59–1.69, p<10^100^) (Figure 1A). A similar pattern of separation by PRS-defined risk in the cumulative incidence of CAD was also seen in all age-by-sex subgroups (Figure 1B), with the pattern seen most clearly in men due to their greater incidence of CAD events.

**Figure 1:**
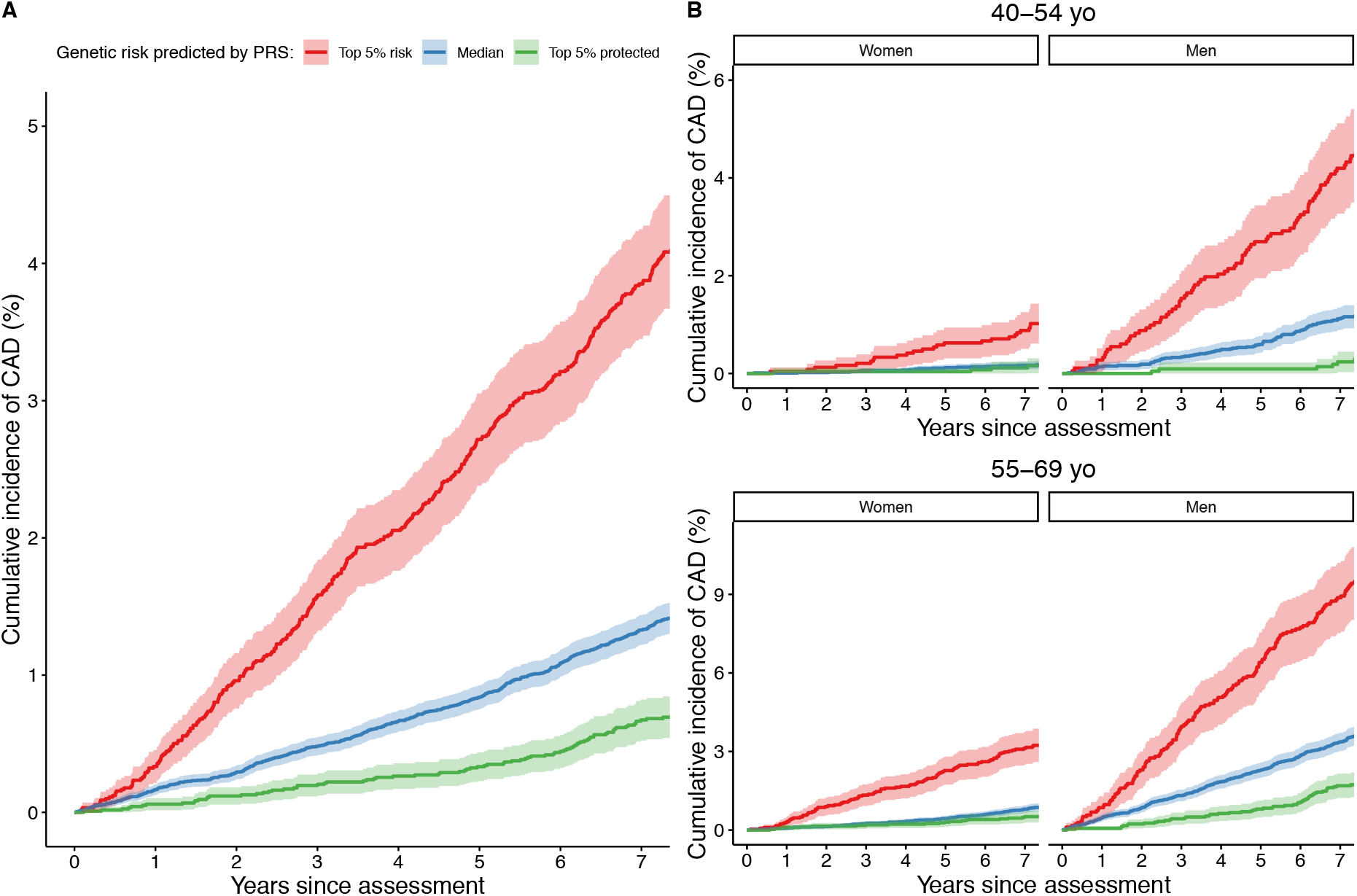
Cumulative incidence of CAD in UKB incident cases in Group III. (A) All ofGroup III. (B) Group III stratified into four subgroups according to age (45–54yo and 55–69yo age ranges) and sex. Individuals are further stratified by PRS-defined risk into the top 5% of PRS risk (red), the median 40–60% distribution of risk (blue), and the bottom 5% of risk distribution (green).

We compared the performance of our PRS against that recently derived and used by Elliott et al ^12^ and also the earlier PRS of Khera et al ^10^ (which was used in the analysis of Mosley et al ^13^). We were unable to conduct a formal comparison in UKB against the PRS developed by Mars et al ^14^, as they used the entirety of UKB to construct their PRS. Published weights for the other two PRS were downloaded and re-applied within our pipeline, in order to provide a like-for-like comparison. Our PRS is the most powerful, followed by the Khera PRS, and then the Elliott PRS (Table 1). When all outcomes are assessed (prevalent and incident outcomes in Groups III and IV), our PRS is significantly better than the second ranking Khera et al PRS (after Bonferroni correction for 4 tests, p = 7.2×10^−11^ for difference in Harrell’s C, p = 4.0×10^−4^ for difference in HR per SD). When only incident outcomes are assessed (Group III), the pattern is similar but the confidence intervals are wider, yielding non-significant results (after Bonferroni correction for 4 tests, p = 0.064 for difference in Harrell’s C, p = 0.44 for difference in HR per SD).

**Table 1:** Prediction performance metrics (with 95% CI) of our PRS compared to that used by Elliott et al ^12^ and Khera et al ^10^. All outcomes (prevalent and incident) were assessed using Groups III and IV. Incident outcomes were assessed using Group III only.

| PRS | All outcomes |  | Incident outcomes |  |
| --- | --- | --- | --- | --- |
|  | Harrell's C | HR per SD | Harrell's C | HR per SD |
| Elliott et al | 0.612 (0.608-0.616) | 1.48 (1.46-1.51) | 0.602 (0.592-0.612) | 1.46 (1.41-1.51) |
| Khera et al | 0.641 (0.637-0.645) | 1.67 (1.64-1.69) | 0.620 (0.610-0.630) | 1.58 (1.53-1.64) |
| This study | 0.660 (0.656-0.664) | 1.74 (1.71-1.77) | 0.637 (0.627-0.647) | 1.64 (1.59-1.70) |

We used generalised survival analysis to examine whether the performance of our PRS varied significantly by age. We found evidence in men, but not in women, that predictive power is highest at younger age groups, and declines for older ages (Figure S2). In contrast, PRS performance (as measured by HR per 1 SD increase) did not significantly differ between men and women (interaction test p-value=0.2 from survival analysis).

We compared the predictive accuracy of our CAD PRS to the individual effects of other known risk factors for CAD. Measured via Harrell’s C, the CAD PRS has similar predictive power to each of systolic blood pressure, HDL cholesterol, and LDL cholesterol, and is more powerful than either total cholesterol or smoking history (Figure S3). The C values vary by age for many of the factors. The only significant difference by sex within age group (at p = 8×10^−4^ after Bonferroni correction for multiple comparisons) is for PRS in the younger (40–55yo) age group. The CAD PRS in 40–55yo men was more predictive than any other single risk factor in any other group.

### Integrated Risk Tool performance

We investigated PCE applied to CAD outcomes as the basis for our primary analysis (see Tables S3-S5 and Figure S4 in the supplementary appendix for secondary analyses relating to a second risk predictor, namely QRISK3, and to a second disease outcome, namely CVD, with qualitatively similar, but less strong, results). We first checked that PCE was a strong predictor in its own right, as previous work had suggested that if PCE is a poor predictor then it becomes an easily beaten “straw man” ^27^. Our overall Harrell’s C for PCE was 0.77 (95% CI 0.76–0.78), comparable to that reported by Elliott et al ^12^ (overall C = 0.76, 95% CI 0.75–0.77), and generally reflective of good prediction ^27^.

We proceeded to assess whether CAD PRS further enhanced risk prediction beyond the PCE predictions. We first calculated the correlation between an individual’s PCE score (log-odds scale) and their CAD PRS, and found that these were largely uncorrelated (Pearson’s correlation coefficient *r* = 0.015; a similar result was found for QRISK3, *r* = 0.025). We note that the two largest contributors to PCE scores are age and sex, which under standard conditions are uncorrelated with PRS. Family history is partly a proxy for genetic risk and is used as a risk factor in QRISK3. However, we again found that our PRS was largely uncorrelated with the family history field in UKB (*r* = 0.084).

We then investigated reclassification patterns, comparing our IRT with PCE and using a 7.5% 10-year risk threshold to define high and low risk groups as recommended under American College of Cardiology / American Heart Association guidelines ^4^ (Table 2). We found substantial reclassification movements between the PCE and IRT models. Overall,13.6% of individuals are reclassified, of which 7.0% are reclassified from low to high risk and 6.6% are reclassified from high to low risk by the IRT. There are also substantial differences by age and sex in the overall number of people that are reclassified by the IRT model, with the overall rate peaking in men at 50–54yo and in women at 65–69yo (Figure S5). Evidence that these reclassifications are beneficial is indicated by the observation that 9.7% of incident cases are correctly up-classified by the IRT, compared to 3.7% that are incorrectly down-classified. Further evidence for beneficial reclassification is provided by a comparison of cumulative CAD incidence in different reclassification groups, which shows that individuals who were up-classified by our IRT had consistently greater cumulative CAD incidence than those down-classified by our IRT (Figure 2). This pattern is also seen to varying degrees in the age-by-sex subgroups (Figure S6).

**Figure 2:**
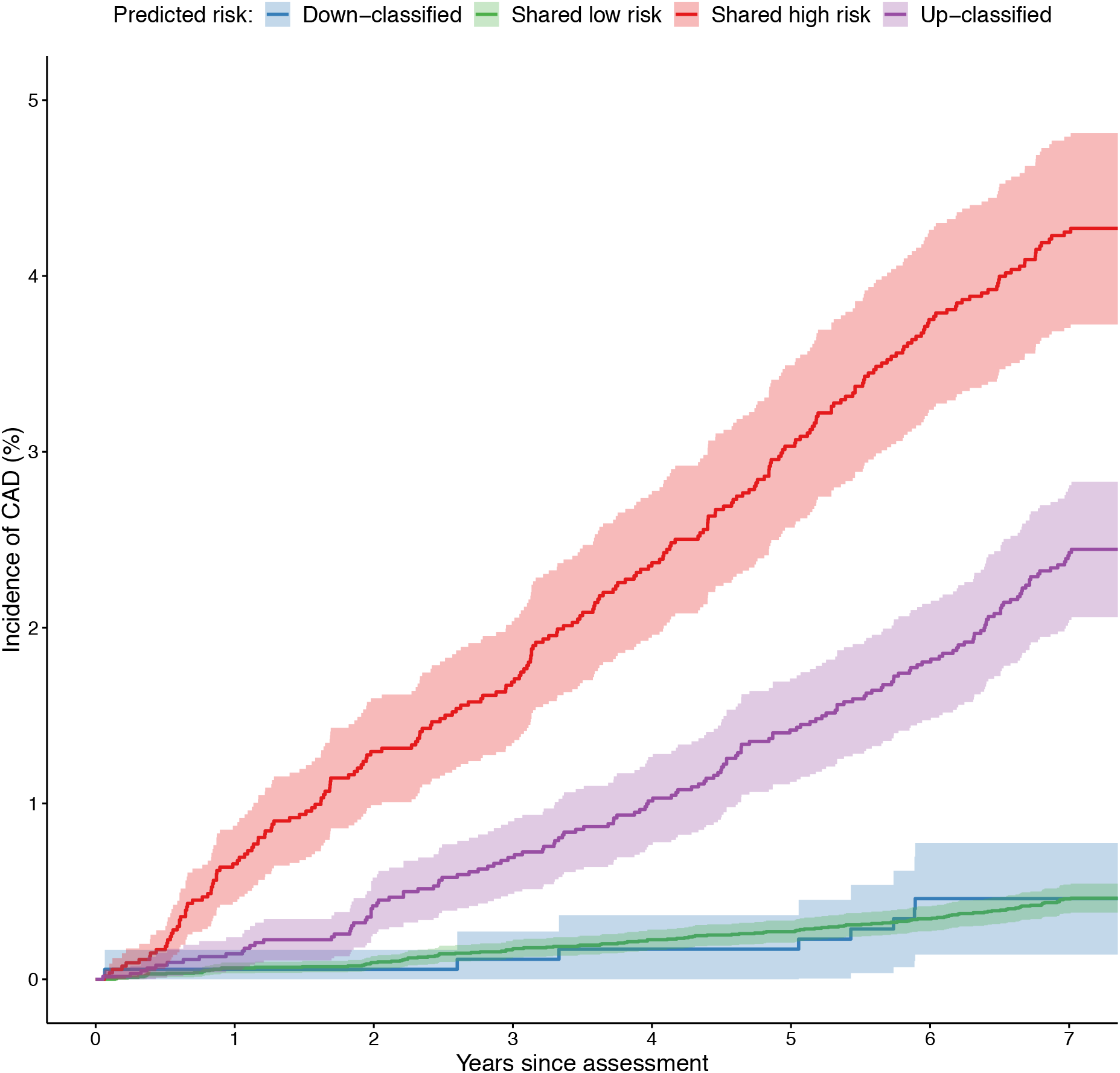
Cumulative incidence of CAD in the subgroup of 40–54yo men in Group III. Individuals are stratified by PCE and IRT-defined risk (above/below 7.5% threshold) into those predicted to be high risk by both PCE and IRT (red), those up-classified to high risk by IRT (purple), those down-classified to low risk by IRT (blue), and those predicted to be at low risk by both PCE and IRT (green).

**Table 2:** Reclassification numbers for our IRT (PCE+PRS) model compared to PCE alone in Group III. Individuals are stratified by sex and age at UKB assessment (40–54yo and 55–69yo). Up/down arrows denote up-/down-classified individuals respectively (predicted to be in a different category by IRT compared to PCE). Horizontal arrows represent individuals predicted to be in the same category by both models. The last column shows the observed number of CAD events in the different reclassification groups, in the 7-year follow-up period post-UKB assessment.

| Sub-group | category | PCE predictions | IRT (PCE+PRS) predictions | Reclassification groups (% of population) | N CAD cases, 7 years (% of total) |
| --- | --- | --- | --- | --- | --- |
| Men, 40-54yo | Above risk threshold | 7,093 (17.92%) | 11,557 (29.2%) | → 5,333 (13.47%) | 226 (44.75%) |
|  |  |  |  | ↑ 6,224 (15.72%) | 151 (29.90%) |
|  | Below risk threshold | 32,490 (82.08%) | 28,026 (70.8%) | ↓ 1,760 (4.45%) | 8 (1.58%) |
|  |  |  |  | → 26,266 (66.36%) | 120 (23.76%) |
| Women, 40-54yo | Above risk threshold | 444 (0.87%) | 992 (1.94%) | → 344 (0.67%) | 14 (8.75%) |
|  |  |  |  | ↑ 648 (1.27%) | 15 (9.38%) |
|  | Below risk threshold | 50,586 (99.13%) | 50,038 (98.06%) | ↓ 100 (0.2%) | 1 (0.63%) |
|  |  |  |  | → 49,938 (97.86%) | 130 (81.25%) |
| Men, 55-69yo | Above risk threshold | 46,861 (91.7%) | 41,663 (81.53%) | → 40,383 (79.02%) | 1,650 (94.12%) |
|  |  |  |  | ↑ 1,280 (2.5%) | 23 (1.31%) |
|  | Below risk threshold | 4,243 (8.3%) | 9,441 (18.47%) | ↓ 6,478 (12.68%) | 67 (3.82%) |
|  |  |  |  | → 2,963 (5.8%) | 13 (0.74%) |
| Women, 55-69yo | Above risk threshold | 23,544 (33.23%) | 24,569 (34.68%) | → 17,817 (25.15%) | 398 (54.15%) |
|  |  |  |  | ↑ 6,752 (9.53%) | 117 (15.92%) |
|  | Below risk threshold | 47,302 (66.77%) | 46,277 (65.32%) | ↓ 5,727 (8.08%) | 41 (5.58%) |
|  |  |  |  | → 40,550 (57.24%) | 179 (24.35%) |
| Overall | Above risk threshold | 77,942 (36.67%) | 78,781 (37.06%) | → 63,877 (30.05%) | 2,288 (72.57%) |
|  |  |  |  | ↑ 14,904 (7.01%) | 306 (9.71%) |
|  | Below risk threshold | 134,621 (63.33%) | 133,782 (62.94%) | ↓ 14,065 (6.62%) | 117 (3.71%) |
|  |  |  |  | → 119,717 (56.32%) | 442 (14.02%) |

Next, we analysed differences in model discrimination and net reclassification improvement (Table 3 and Figure 3). Overall, the difference in Harrell’s C was 3% (95% CI 2%−4%), and ranged from 1%−5% in the age-by-sex subgroups. The overall NRI was 5.7% (95% CI 4.4%− 6.0%). The positive changes in full NRI were all strongly significant even after Bonferroni correction for multiple testing (maximum p = 0.005 after correcting for the overall test plus four age-by-sex tests), while the difference in Harrell’s C was significant overall (corrected p = 4.0×10^−7^) and in the two male subgroups (men 40–54yo corrected p = 5.0×10^−4^; men 55–69yo corrected p = 1.3×10^−6^) but not in the two female subgroups. Broadly similar results were obtained when the right-censoring of CAD outcomes was moved from 7 years to 10 years in both training and test sets (Table S6). Stated in traditional diagnostic terminology (sensitivity, specificity, and positive predictive value) at the established 7.5% 10-year risk threshold, our rescaling approach holds the positive predictive value constant while improving sensitivity at a moderate specificity cost (Table S7). The parameters of this trade-off vary depending on the subset of the population being considered (Table S7).

**Table 3:** Prediction performance metrics (with 95% CI) for incident CAD outcomes in Group III, comparing PCE and IRT (PCE+PRS) models and stratifying into age-by-sex subgroups.

| category | Harrell's C |  |  | NRI (PCE vs IRT) |  |  |
| --- | --- | --- | --- | --- | --- | --- |
|  | PCE | IRT | Difference | Full | Within-cases | Within-noncases |
| Overall | 0.77 (0.76–0.78) | 0.80 (0.79–0.81) | 0.03 (0.02–0.04) | 5.68 (4.41–6.96) | 5.99 (4.73–7.26) | -0.31 (-0.47 – -0.15) |
| Men (40-54yo) | 0.75 (0.73–0.77) | 0.80 (0.78–0.82) | 0.05 (0.02–0.08) | 17.26 (13.01–21.51) | 28.32 (24.09–32.54) | -11.06 (-11.49 – -10.63) |
| Women (40-54yo) | 0.75 (0.71–0.79) | 0.76 (0.72–0.80) | 0.01 (-0.05–0.07) | 7.70 (2.99–12.41) | 8.75 (4.04–13.46) | -1.05 (-1.15 – -0.95) |
| Men (55-69yo) | 0.65 (0.63–0.66) | 0.69 (0.68–0.71) | 0.05 (0.03–0.07) | 8.18 (7.01–9.35) | -2.66 (-3.78 – -1.54) | 10.84 (10.49–11.19) |
| Women (55-69yo) | 0.68 (0.66–0.70) | 0.72 (0.70–0.74) | 0.04 (0.01–0.07) | 9.67 (6.08–13.26) | 11.75 (8.17–15.32) | -2.07 (-2.39 – -1.76) |

**Figure 3:**
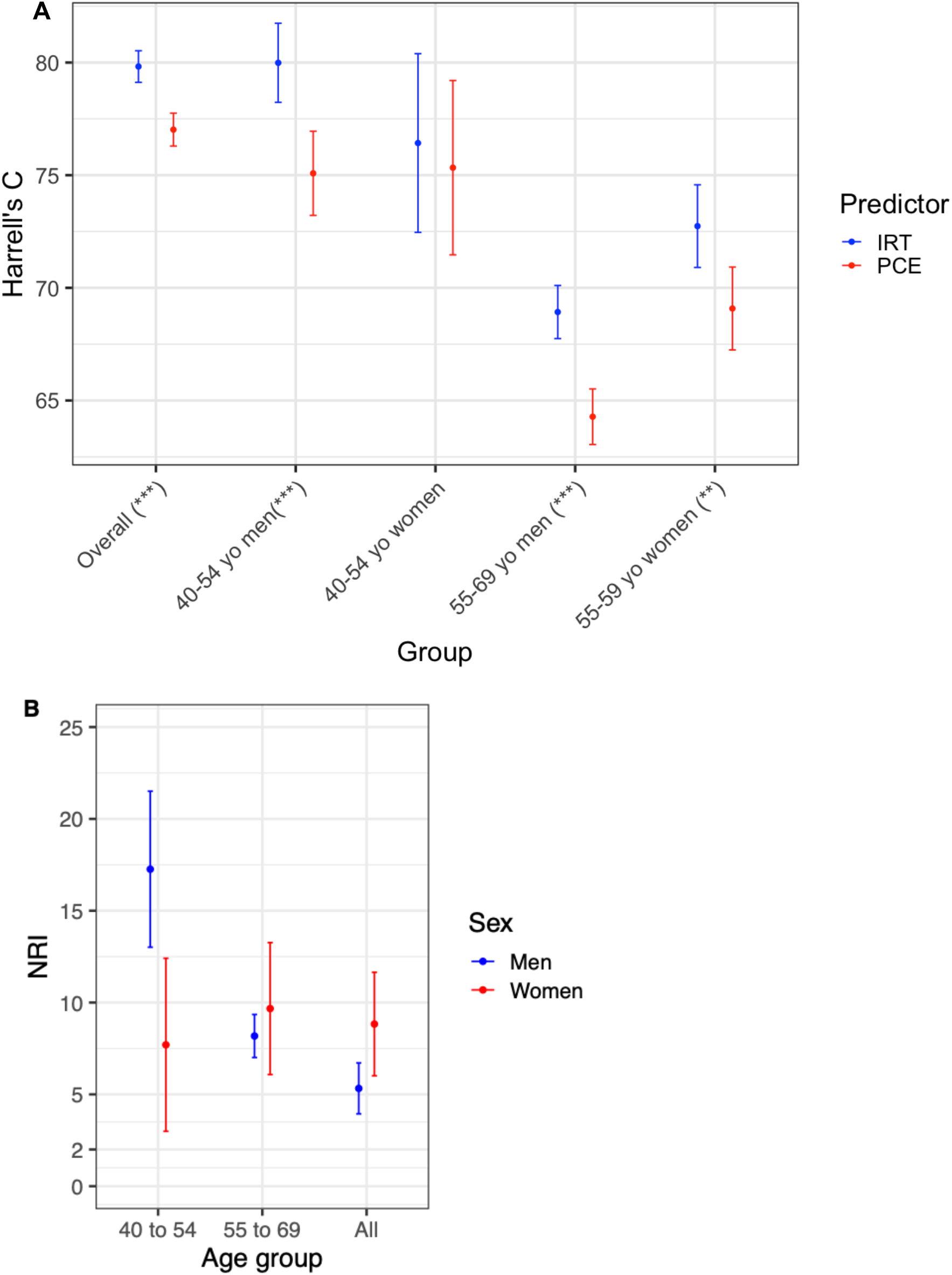
Model discrimination and net reclassification improvement for the IRT compared to PCE. (A) Harrell’s C overall and across age-by-sex subgroups. Blue and red lines refer to IRT and PCE respectively. (B) Net reclassification improvement (NRI) for the IRT compared to PCE alone across different age groups in men (blue) and women (red). The bars indicate the 95% CIs.

When broken down into age-by-sex subgroups, we observed that all the subgroup NRIs were larger than the overall NRI, ranging from 7.7%−17.3% and with the largest improvement seen in younger middle-aged men (40–54yo) (Table 3). This behaviour is driven by different types of positive reclassification in younger (40–54yo) versus older (55–69yo) middle-aged men, which to some extent cancel out in the overall NRI. In younger men, the large positive NRI (17.3%, 95% CI 13.0%−21.5%) is driven by a very large net reclassification improvement in cases (28.3%, 95% CI 24.1%−32.5%), while in older men the positive NRI (8.2%, 95% CI 7.0%−9.4%) is driven by a positive net reclassification improvement in noncases (10.8%, 95% CI 10.5%−11.2%) (Figure 3B and Figure S7).

## Discussion

In our study, we developed a new PRS for CAD, and then considered its predictive performance both on its own and when used as part of an integrated risk tool (IRT) by combining the CAD PRS with established clinical risk prediction tools based on non-genetic risk factors (PCE and QRISK3 as recommended in the US and UK respectively). We evaluated performance of the IRT in a large independent test set of 212,563 participants in the UK Biobank (UKB), both overall and in participants stratified into age-by-sex subgroups.

In contrast to previous studies ^12–14^, we found that our IRT performed substantially better than the established risk prediction tools. Previous studies reported overall net reclassification improvements in the range 0.1% – 4.4%. We report an overall net reclassification improvement of 5.7% compared to PCE. This numerical increase translates to substantive clinical utility, as we discuss below.

There are several ways in which our study differs from the other three cited above, but we highlight two that appear to have had a substantial impact. Firstly, we developed and applied our own distinct PRS construction method for CAD, in which we combined the largest available published CAD GWAS with additional UKB data, leveraged cross-trait genetic pleiotropy in > 5,000 other GWAS to aid in the identification of CAD associated loci, and used functional information to aid in the calculation of optimal PRS weights. Our novel PRS provides more powerful prediction than the PRS published by Khera et al ^10^ (and used by Mosley et al ^13^). It is also substantially more powerful than the PRS developed by Elliott et al ^12^, which may explain some of the differences between our results and those of the latter study. When our CAD PRS was considered as a risk factor on its own, we found it to be as powerful for prediction as each of several established measured risk factors (systolic blood pressure, HDL, and LDL cholesterol), and more powerful than others (e.g. total cholesterol and smoking history). It is the also best performing single risk factor in younger middle-aged men (40–54yo).

Secondly, we analysed in detail the break-down of reclassification losses and gains making up the net reclassification improvement. Net classification improvements within age-by-sex subgroups are all higher than the overall figure (ranging from 7.7% – 17.3%), with a particularly striking value of 17.3% for younger middle-aged men (40–54yo). A net additional 28.3% of cases are identified by our IRT in this subgroup that are overlooked by the PCE tool. We note that observed cases in our study are from a 7-year follow-up window, which means some of our observed noncases will in fact be cases occurring between 7 to 10 years, and this in turn means our reclassification statistics for cases are likely to be more accurate than those for noncases.

There are limitations to our study, most of which are shared by other PRS studies. Our study was performed in the UK Biobank, and is therefore limited by the characteristics of this cohort. In particular, the cohort is of primarily European ancestry (and was restricted to European ancestry in this study), the age range of participants at UKB assessment is limited to 40–69yo, and participants tend to be healthier and more affluent than the general UK population ^28^. As previously noted, the 7-year follow-up time post UKB assessment was less than the 10-year risk period addressed by both the PCE and QRISK3 algorithms, leading to some false labelling of noncases as cases. This means that, for all the models assessed in this study, both those with and those without a PRS, case misclassifications were overestimated and noncase misclassifications were underestimated. Additional limitations include the assessment of a simple single-risk-assessment-at-baseline scenario rather than a continuous assessment scenario, a blanket exclusion of samples with missing data for training and testing, a PRS that is constructed from common variants without rare high-risk variants, an evaluation of PCE and QRISK3 risk tools only, a reliance on UKB data that included self-report to define some outcomes and variables, and the use of prevalent CAD cases in the construction of the PRS.

An additional limitation of our study, which is common to all PRS work so far, is that we define a single PRS and apply it uniformly to all individuals, regardless of their age, sex, or other relevant factors. This approach assumes that, throughout the genome, common variant genetic effects are independent of these other factors. In contrast, we have shown here that PRS prediction varies by age in men. This opens the possibility that once sufficient data are available, separate PRSs may be constructed for different groups (defined for example by age, sex, or ancestry background) which will likely further improve predictive power ^29^. Even without these more sophisticated statistical approaches, the predictive power of PRSs will also increase as more population-scale data becomes available.

One potential concern with using PRS to identify high-risk individuals is whether existing interventions are effective in genetically defined risk groups. Studies performed so far are reassuring in this regard. Statin therapy ^30,31^ and PCSK9 inhibition ^32^ is at least as effective in individuals with high CAD PRS, and may in fact be more effective than average. Lifestyle changes involving diet and exercise are also effective in this group ^33^.Cardiovascular risk tools have continually evolved as additional risk factors have been shown to improve a tool’s predictive ability, as for example with the addition of diabetes information that was not part of the original Framingham risk score ^3,34^. It has long been known that CAD has a large heritable component ^7,8^. Our analyses describe the most powerful genetic predictor to date for the polygenic component of CAD risk. The predictive accuracy of our PRS alone is comparable to most current clinical risk factors captured in the PCE, and is better than some. An advantage of PRS as a risk factor is that it can also be used earlier in an individual’s life to identify those who may have high lifetime CAD risk, but before most other non-genetic risk factors have predictive power. We further show that the new CAD PRS is uncorrelated with PCE and QRISK3, suggesting that its combination with these risk tools should offer enhanced risk prediction. Family history (not formally part of PCE, but part of QRISK3) is an imperfect proxy for genetic risk, as well as for shared environmental risk factors, and our study shows a low correlation (*r* = 0.08) between family history and our PRS.

In the US, cardiovascular disease risk assessment is recommended in people aged 40 to 69 years ^4^, representing 37% of the US population (∼122 million individuals). The addition of our PRS to PCE would up-classify 7% of the population to a level of cardiovascular risk that warrants statin prevention (∼8.5 million individuals). If these participants were offered statin therapy, and assuming a benefit of 250 events avoided over 5 years per 10,000 individuals ^35^, ∼210,000 major cardiovascular events would be avoided. Assuming a 10% mortality rate from a major cardiovascular event ^36^, this prevention strategy would imply ∼21,000 deaths avoided in the US over a 5 year period.

We therefore conclude that the addition of the polygenic risk score enhances the predictive ability and clinical utility of existing CAD risk tools. This enhanced predictive ability is especially pronounced in younger middle-aged men (40–54yo), but it is seen across all studied age groups and across sexes. Our study argues that future iterations of PCE (and similar tools) may benefit from the addition of polygenic risk scores.

## Data Availability

The values of the polygenic risk scores using our novel CAD PRS for the 212,563 individuals on whom they were evaluated in the paper will be returned to UK Biobank so that they can be made available to approved UK Biobank researchers. The UKB Group I GWAS summary statistics will be shared by Genomics plc on a public data repository. Please contact author VP for details.

## Contributors

VP and PD conceived and led the study. FRM analysed the data and generated the figures and tables. MEW, FRM, VP, PD led the write-up of the study. RM, EK, RMS, SS, WAT, PS, JAG, AS, CCAS developed the analytics that enabled the derivation of the PRS, the UKB phenotype inference and the evaluation of the models in UK Biobank. JH-C, DH, ASL, JWOS, EAA, JD commented and edited the manuscript.

## Acknowledgements

We thank all the participants, scientists, and funders who generated and shared the data from the UK Biobank and other data sources that were used in this study. GWAS data on coronary artery disease have been contributed by CARDIoGRAMplusC4D investigators and have been downloaded from http://www.cardiogramplusc4d.org.

## Supplementary figures

**Figure S1:** Flowchart defining the UK Biobank sub-cohorts used in this study.

**Figure S2:** Age dependent survival analysis in Group III. (A) All of Group III. (B) Group III stratified into two subgroups according to sex. The lines show the estimated CAD hazard ratio (shaded area: 95% CI) as a function of age at diagnosis for individuals in the top 10% of the CAD PRS distribution compared to the median of the population.

**Figure S3:** Harrell’s C for some of the risk factors that contribute to CAD risk, based on a Cox regression analysis on CAD outcome in Group III (incident cases only) and stratified by both age and sex. Men and women are represented by blue and red lines respectively. 40–54yo and 55–69yo age groups are represented by dotted and dashed lines respectively. Overall Harrell’s C is represented by a solid black line. Error bars indicate the 95% CI.

**Figure S4:** Combined (A), within-case (B) and within-noncase (C) net reclassification improvement (NRI) for our integrated risk tool (PRS+QRISK3) compared to QRISK3, using individuals in Group III and stratified by age and sex and using a 10% risk threshold to define high/low risk individuals.

**Figure S5**: Percentage of the population reclassified by adding our integrated risk tool to PCE. The solid line shows the overall percentage of reclassification, and the dashed line shows the proportion of individuals up-classified. Blue and red lines refer to men and women respectively.

**Figure S6:** Cumulative incidence of CAD in Group III individuals stratified by age (panel A for 40–54yo and panel B for 55–69yo) and sex (subpanels within A and B). Individuals are further stratified by PCE and IRT-defined risk (above/below 7.5% threshold) into those predicted to be high risk by both PCE and IRT (red), those up-classified to high risk by IRT (purple), those down-classified to low risk by IRT (blue), and those predicted to be at low risk by both PCE and IRT (green).

**Figure S7:** Within-case (A) and within-noncase (B) net reclassification improvement (NRI) for our integrated risk tool (PRS+PCE) compared to PCE, using individuals in Group III and stratified by age and sex and using a 7.5% risk threshold to define high/low risk individuals.

